# Whole-Genome Sequencing Of Omicron Identified Multiple Outbreaks And Introduction Events In India During November 2021 and January 2022

**DOI:** 10.1101/2022.04.20.22270880

**Authors:** Shreedhanya D Marathe, Varun Shamanna, Geetha Nagaraj, S Nischita, Muthumeenakshi Bhaskaran, K L Ravikumar

## Abstract

The variant of Concern(VOC), Omicron is the predominant variant circulating throughout the world of the SARS COV2 pandemic during the third wave including India. The World Health Organisation has designated this highly mutated variant as a VOC due to its high transmissibility and risk of reinfection. Whole-genome sequencing and analysis were performed for SARS-CoV2 PCR positive samples between Dec21 to Jan22. From the 133 omicron variants detected, genomic analysis was carried out by contextualizing them with 1586 complete genomes of Omicron from India obtained from GISAID.

The Omicron variant prevalence in India has increased in a log phase within 3 months in most of the metropolitan cities. The sublineage BA.1 was first observed in the country, while the BA.2 sublineage was introduced to Delhi in the mid of December 2021. The two outbreaks observed were of BA.2 variant and were observed to spread to multiple cities in a short time. The rapid spread and specific mutations in the outbreak samples of Omicron indicate that the variant is highly transmissible when compared to previous variants. The study shows the importance of genomic sequence to identify the emergence of clusters and take actions to prevent further spreading events.

## INTRODUCTION

Coronavirus is a virus that infects the nose, sinuses, and upper respiratory tract, most of them not causing severe effects [1]. After an outbreak in Wuhan, China in December 2019, the World Health Organisation (WHO) recognized severe acute respiratory syndrome coronavirus 2 (SARS-CoV2) as a novel coronavirus that was a serious threat to humans [2]. The SARS-CoV2 virus, like other coronaviruses, has been undergoing continuous mutations, resulting in multiple variants of concern (VOC) [3].

On November 25, 2021, the WHO identified the Omicron (B.1.1.529) variation of SARS-CoV-2 to be a variant of concern after its initial identification in Gauteng Province, South Africa. The Omicron variant’s rapid spread sparked the fourth wave of SARS-CoV-2 infections in South Africa, with daily diagnosed cases exceeding totals reported in the country during all preceding periods [4]. Various reports from across the globe have shown that there is a significant increase in the spread of the omicron variant when compared to the delta [5, 6]. As of January 2022, the Omicron variant has disseminated worldwide including India, where 90% of all SARS-CoV-2 infections diagnosed in the metro cities in January 2022 were estimated to be caused by the Omicron variant [32]. The rise in the positivity rate resulted in the declaration of a third-wave crisis in the country [7]. The spread of SARS-CoV2 widely and causing a high risk of infection has led to a greater opportunity for mutation and possible positive evolution [4]. This variant is known to contain numerous mutations in the RBD of the spike protein which has led to its maximum transmissibility. It has been hypothesized that omicron would have emerged from Alpha that had stemmed from a persistently infected patient who got infected with multiple variants. But as the number of cases is increasing and mutation analysis is becoming more robust, we are deviating from this hypothesis and concluding that omicron is emerging with multiple mutations from different variants [8]. At least 50 mutations differentiate the Omicron variant from the SARS-CoV-2 reference strain. The spike protein alone possesses 30 of the mutations with some overlapping mutations with previously identified strains as well as mutations specific to omicron. Preliminary laboratory studies have shown that Omicron can escape extensively, but incompletely, from neutralizing antibodies in vaccinated and convalescent sera [9].

Covid 19 pandemic is the first where real-time Whole Genome Sequencing (WGS) was performed to track transmission, identify vaccine targets, and design therapeutics for the virus. The accurate identification and sequencing of SARS-CoV-2 in patient samples provides extensive information, ranging from reducing false negatives and contact tracing to evaluating the suitability of diagnostic assays (particularly nucleic acid-based) and determining whether vaccines are likely to be or remain effective despite genetic recombinations, diverse mutations during virus replication [10].

We performed and analyzed WGS of SARS-CoV2 samples sequenced as a part of INSACOG and compared with existing genome sequences of omicron variant available in GISAID from India to study the following:

A. Geo-spatial distribution
B. Demographic characterization
C. Sub lineage proportions
D. SNP characterization

## METHODS

### 2.1 Sequencing of SARS-CoV2 PCR positive isolates

#### 2.1a Clinical specimens

The study was conducted after obtaining ethical approval vide letter number KIMS/IEC/A012/M/2022. Central Research Laboratory, is an ICMR recognized COVID19 testing laboratory set up in Kempegowda Institute of Medical Sciences, a tertiary care hospital and a postgraduate medical college situated in the heart of the Bengaluru city. Nasopharyngeal swabs collected from 01 December 2021 up to the 10th of January 2022 as a part of routine diagnostics were tested for COVID-19 using the ICMR-approved RT-PCR test. The positive samples with a CT Value between 20 to 30 were taken for sequencing as a part of the National SARS COV2 surveillance program INSACOG. The epidemiological metadata was obtained from the SRF forms submitted during the collection of the sample.

#### 2.1b Genome Sequencing

The 192 samples positive for COVID-19 from RT-PCR were sequenced on the nanopore MK1C platform. Initially, 1200 bp amplicons were generated using Midnight PCR primers by doing as per the manufacturer’s specification [9]. The concentrations of cDNA were determined using the dsDNA HS Assay Kit with Qubit 4 Fluorometer. RBK-110-96 rapid barcoding kit [11] was used to barcode each sample and the sequencing was performed on MIN-106 flowcells.

#### 2.1c Sequence analysis

Guppy 3.6.0 was used for base calling and demultiplexing the samples with a 450 bp fast base calling configuration [12]. The demultiplexed reads were assembled using nf-core viralrecon pipeline [13] with medaka g303 model for read-polishing [14]. A minimum of 250bp and a maximum of 1400 bp were used as read-cutoff as per the standards used for midnight protocol. The consensus assembly obtained was used further for variant calling and lineage analysis.

### 2.2 Retrieval of Genomic Data from the database

From the Global Initiative for Sharing all Influenza Data (Gisaid) [15], we downloaded 1586 high coverage, complete VOC Omicron GRA(B.1.1.529+BA.*) SARS-CoV-2 genome sequences submitted from different centres across India as of 19th January 2022. Also, the patient information and metadata for the downloaded sequences were retrieved. The sequences with large gaps and more than 50 ambiguous bases were removed. The sequence of the established SARSCoV-2 reference genome (NC_045512.2; Wuhan, December 2019) was downloaded from NCBI for further variant analysis and gene annotations.

### 2.3 SNP identification and Phylogenetic analysis

#### 2.3a SNP identification and Lineage prediction

The consensus sequence files from in-house sequence analysis and those downloaded from GISAID were combined and were submitted to https://clades.nextstrain.org/ to get the type defining markers and clade assignment. Nextclade, the open-source web-based tool provided Mutation detection, clade assignment, quality checks, and phylogenetic placement of the isolates [16]. The genomic dataset was classified into sub-lineages using PANGOLIN (Phylogenetic Assignment of Named Global Outbreak LINeages) [17].

#### 2.3b Phylogenetic tree construction

The consensus sequences were aligned to the reference genome to create multiple sequence alignment using MAFFT [18] and the alignment positions tagged as inappropriate for phylogenetic analysis were removed. Maximum likelihood phylogenetic reconstruction was performed with the curated alignment and a General Time Reversible (GTR) model using FASTTREE with 1000 bootstrap replicates [19].

### 2.4 Visualisation of the tree

The microreact [20] was used to view the tree & analyze the SNPs and distribution of the sub-lineages across various states in the span of 3 months i.e, November 2021, December 2021, and January 2022.

## RESULTS

### 3.1 Genomic data

A total of 3429 in-house samples were tested on RT-PCR for COVID-19 from December 2021 to January 2022 from Bengaluru, Karnataka. Among them, 235 samples were tested positive and 192 samples that had CT values between 20 to 30 were taken for sequencing. The sequence data provided even genome coverage with high-quality viral genomes to be reconstructed at 500x average depth coverage with an average of 0.1 million reads. Out of 192, the Pangolin tool classified 133 isolates as omicron variants. In addition, 1586 isolates corresponding to the omicron variant are included from GISAID. Thus for further analysis, we included 1719 isolates. The metadata and variant details are provided in supplementary table 1.

### 3.2 Geospatial distribution

On the 10th of November 2021, the first Omicron isolate was collected and tested from Mumbai, Maharashtra. By the time WHO declared Omicron as the VOC, it had already spread to multiple states like Delhi, Kerala, Gujarat, and Karnataka. As shown in Fig 1a maximum sequences were uploaded in the month of Dec 2021 out of which 29% were classified as Omicron. Then in the month of Jan up to 19th 2022, 84% of the total sequences were classified as Omicron. We can observe the takeover of the omicron variant with respect to other variants. By the beginning of January 2022, the virus had extended its reach to further states like Telangana, Punjab, Andhra Pradesh, etc, and community transmission was seen and the state-wise distribution is shown in Fig 1b.

**Fig 1a:**
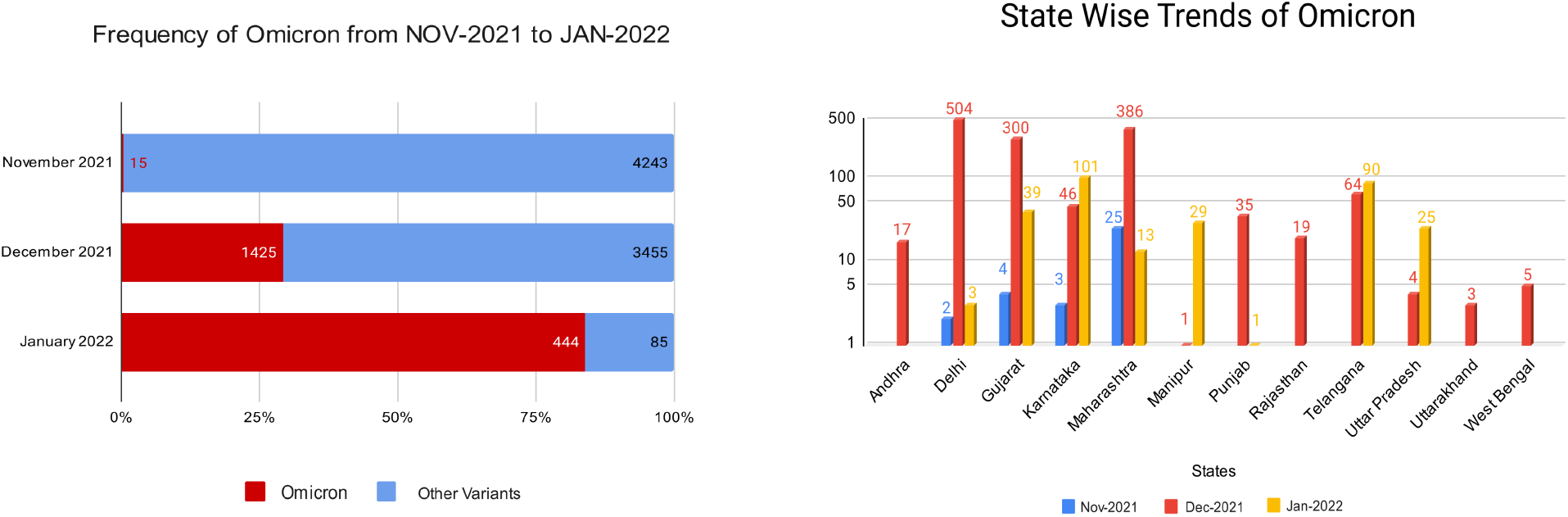
Number of samples in different months. The bar represents the total samples in that month and Red color shows the percentage of samples belonging to the omicron variant Fig1b: Number of Omicron samples distributed across different states of India. The height of the bar is proportionate to the number of samples.

### 3.3 Lineage prevalence

Pango lineage B.1.1.529 was the first to be declared as the Omicron VOC. Due to acquisitions of mutations, as it spread globally, sub-lineages emerged as BA.1, BA.2, and BA.3 [21].

Until the mid of December 2021, we observed the prevalence of BA.1 sublineage along with B.1.1.529 in different states of India as shown in Fig 2. Karnataka and Delhi were the first states to report this sublineage in the mid of November 2021.

**Fig 2:**
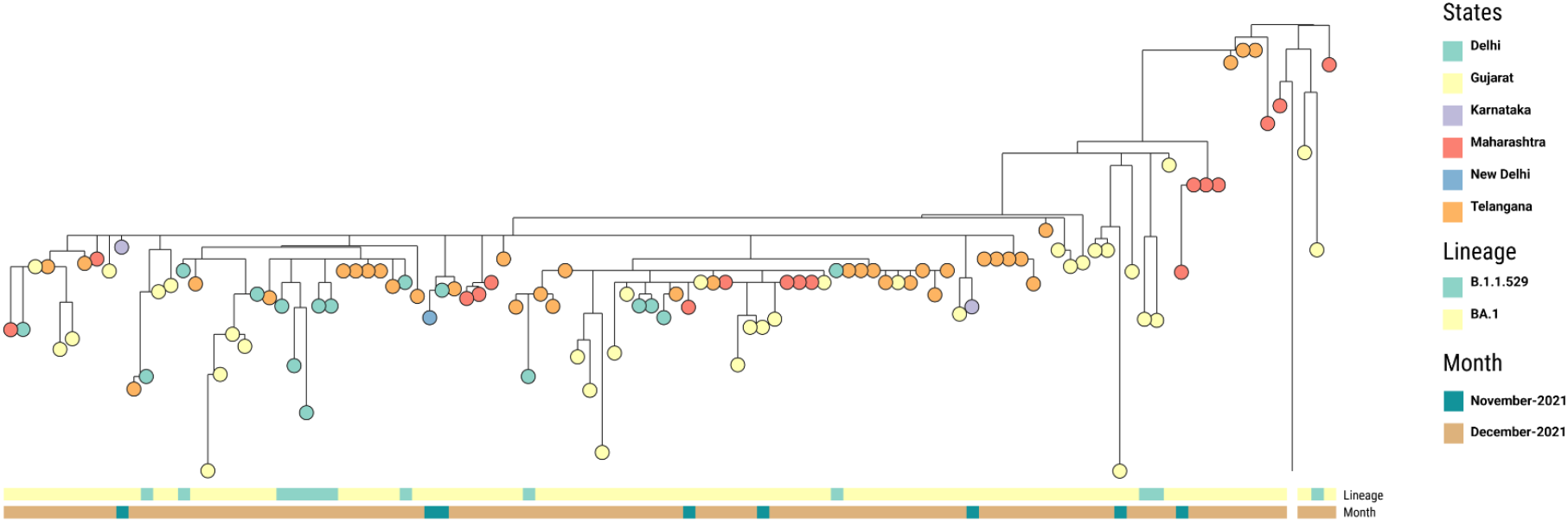
Phylogenetic tree of Omicron sequences between Nov 2021 to Dec 15th, 2021 depicting the prevalence of BA.1 in different states of India. The nodes are colored by states and the metadata block represents the lineages. Can be viewed at https://microreact.org/project/6hU3Rp3Wf8yDK24E2mBvh5-omicronindiafirst

The first case of BA.2 sublineage was found in Delhi on the 16th of December with a cluster of 22 from a single area. We analyzed the SNP difference within this cluster and found that they had 0-1 SNP, which confirmed it could be a potential outbreak. BA.2 was observed in other states like Gujarat, Telangana, Maharashtra, etc with an incidence of 18% (n/total= 312/1719) by December end. N:S413R, S:A27S, S:V213G, ORF1a:L3027F nonsynonymous mutations were specific to the outbreak. However, it was found in less than 30% of the total isolates from India.

We observed another cluster from multiple states with an SNP difference of 0-1. The first case in this cluster was from Maharashtra which originated on the 20th of Dec 2021 and spread to Karnataka and Punjab simultaneously. Out of 51 samples in the cluster, 32 of them were from Bengaluru which were our In-house samples. The remaining 18 isolates belonged to Punjab where it was observed after 4 days of the first detection. This could potentially mean that there were multiple introductions to Bengaluru and Punjab from other metropolitan cities that drove the BA.2 spread. SNP analysis showed that the predominant deletions such as N:E31-, N:R32-, N:S33-, ORF9b:E27-, ORF9b:N28-, ORF9b:A29- and substitutions N:P13L, ORF9b:P10S were detected only in 2% (1/51) and 17.6% (9/51) respectively.

**Fig 3a:**
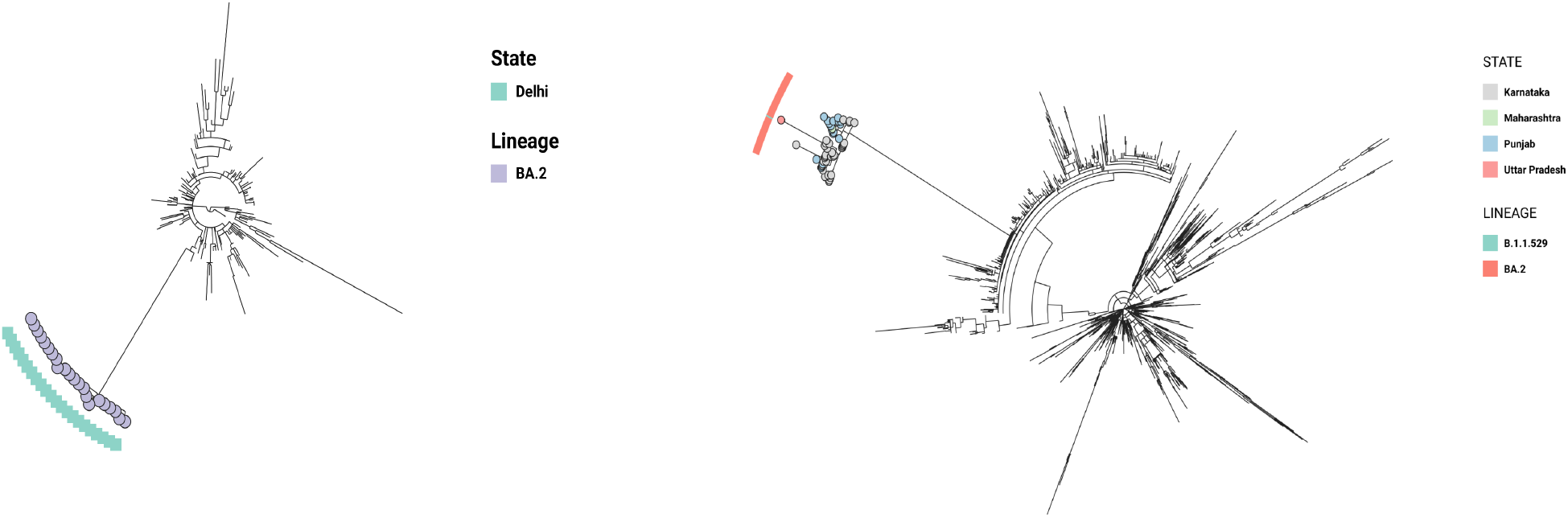
Phylogenetic tree showing the outbreak of BA.2 sublineage in Delhi. The view is available at https://microreact.org/project/2H1zuFXnnAuwvhREBJrW7L-omicronfirstoutbreak Fig3b: Tree showing multistate outbreak of BA.2 variant, the first blocks showing variant and the second blocks showing the states. The view is available at https://microreact.org/project/ankEHTJjr93qMegr4AfJTD-omicronindia

## DISCUSSION

Omicron has quickly spread to become the world’s dominant variant of SARS-CoV2 by the end of December 2021. The Omicron variant belongs to PANGO lineage B.1.1.529, in comparison to the original SARS-CoV-2 strain, this has 30 amino acid modifications, three modest deletions, and one short insertion in the spike protein. In addition to these mutations, a number of substitutions and deletions in other genomic areas are also present in Omicron [28]. Our study showed the presence of specific mutations in the outbreak isolates and the microevolution of the sublineages. The B.1.1.529 lineage was given the name Omicron after it was designated as a variant of concern. Later, it was discovered that this lineage had its own small variations, and the three most common were designated as B.1.1.529.1, B.1.1.529.2, B.1.1.529.3, or simply known as BA.1, BA.2, and BA.3 [29]. In the month of January, BA.1 is the prevalent Omicron sub-variant in circulation while there is a significant rise in the proportion of BA.2 simultaneously.

According to the WHO, >98 percent of all Omicron genetic sequences submitted in global databases till January 15 were that of the BA.1 variant [31]. BA.2 holds most of the characteristics of BA.1 but has some additional mutations that can give it a distinguishing feature. Some of the important mutations in BA.2 that are different from other sub-lineages include the spike protein mutations such as T19I, 24-26 deletion, V213G, T376A, R408S, etc [30]. BA.2 has been discovered in 49 countries, according to the outbreak.info website, which tracks the prevalence of different lineages of this virus around the world [27]. Since the first sequence was available to the public on the 10th of November 2021, we have seen varied proportions of sublineages circulating in India. As seen in the global trend, India also follows the same route.

Omicron was introduced to India in early November 2021. Reports say that it was first seen in Karnataka [22, 23] but the first sequence collected and submitted to GISAID was on the 10th of Nov 2021 from Maharashtra. At the beginning of December 2021, sequences of Omicron were 1% of the total sequences from India. Within a month the Omicron replaced all other variants to become 98% of total sequences. At present 100% of the cases are due to Omicron in India [24]. Even though the reports in early Jan 2022 say that Omicron is in the community transmission stage [25], the data in GISAID shows that the community transmission was seen in mid of Dec 2021 and was a dominant variant by end of Dec 2021 [26]. Even though in the month of Dec 2021 the number of omicron cases remained less than other variants it had already extended to most of the cities in India signifying the community transmission. While it took a month to increase its frequency to 50% of the total cases in the African continent, Europe, Asia, and the American continent saw 75% of the total cases reported as Omicron in the month of December 2021.

We have BA.1 prevailing in different states with 61% (n=1046/1719) of the total cases analyzed. Following this, BA.2 (n=545/1719) was seen in 37% of the cases. At different time frames, we have noticed the introduction of sublineages in different parts of the country. The first outbreak cluster involved BA.2 introduction in the mid of Dec 2021 which then spread across different states within a few days. We found that both the outbreak clusters were caused by the BA.2 sub-lineage. It indicates that BA.2 is highly variable when compared to BA.1. This is causing higher transmission leading to the community spread of Omicron. During the analysis period, BA.3 sublineage was not found in India.

## CONCLUSION

The study provides an in-depth view of the spread and transmission of the omicron variant in India and showed the community transmission had begun in the mid of December 2021 itself. WGS significantly enhanced understanding of SARS-CoV-2 omicron variant transmission and dynamics of outbreaks, identifying spatial and mutational variation of the same variant and within sublineages. Examination of the genomic epidemiology of Omicron variant across India showed the multiple introduction events at different time spans which led to the 3rd wave of the COVID-19 pandemic.

## Data Availability

All supporting data, code and protocols have been provided within the article or through supplementary data files.

https://microreact.org/project/6hU3Rp3Wf8yDK24E2mBvh5-omicronindiafirst

https://microreact.org/project/2H1zuFXnnAuwvhREBJrW7L-omicronfirstoutbreak

https://microreact.org/project/ankEHTJjr93qMegr4AfJTD-omicronindia

## ACKNOWLEDGMENTS

The work was carried out at the Central Research Laboratory, Kempegowda Medical College, Bengaluru as a part of The Indian SARS-CoV-2 Genomics Consortium (INSACOG).

## Notes

**CONFLICT OF INTERESTS:** The authors declare that there is no conflict of interest.

### Competing Interest Statement

The authors have declared no competing interest.

### Funding Statement

The authors declare no funding was received for the study.

### Author Declarations

The study was approved by the Kempegowda Institute of Medical Science independent ethics committee (Approval ID: KIMS/IEC/A012/M/2022). The study was conducted according to the guidelines and recommendations of Good Clinical Practice and the Declaration of Helsinki. Written informed consent was obtained from each participant or legal guardian as applicable.

